# Increased Transmissibility of SARS-CoV-2 Lineage B.1.1.7 by Age and Viral Load: Evidence from Danish Households

**DOI:** 10.1101/2021.04.16.21255459

**Authors:** Frederik Plesner Lyngse, Kåre Mølbak, Robert Leo Skov, Lasse Engbo Christiansen, Laust Hvas Mortensen, Mads Albertsen, Camilla Holten Møller, Tyra Grove Krause, Morten Rasmussen, Thomas Yssing Michaelsen, Marianne Voldstedlund, Jannik Fonager, Nina Steenhard, The Danish Covid-19 Genome Consortium, Carsten Kirkeby

## Abstract

1

**Aim:** The aim of this study was to estimate the household transmissibility of SARS-CoV-2 for lineage B.1.1.7 compared with other lineages, by age and viral load. Furthermore, we wanted to estimate whether there is a multiplicative or additive effect of the increased transmissibility of B.1.1.7 compared with other lineages.

**Background:** New lineages of SARS-CoV-2 are of potential concern due to higher transmissibility, risk of severe outcomes, and/or escape from neutralizing antibodies. Lineage B.1.1.7 has been estimated to be more transmissible than other previously known lineages, but the association between transmissibility and risk factors, such as age of primary case and viral load is still unknown.

**Methods:** We used comprehensive administrative data from Denmark, comprising the full population, all SARS-CoV-2 RT-PCR tests, and all WGS lineage data (January 11 to February 7, 2021), to estimate household transmissibility stratified by lineage B.1.1.7 and other lineages.

**Results:** We included 5,241 households with primary cases; 808 were infected with SARS-CoV-2 lineage B.1.1.7 and 4,433 were infected with other lineages. The attack rate was 38% in households with a primary case infected with B.1.1.7 and 27% in households with a primary case infected with other lineages. Primary cases infected with B.1.1.7 had an increased transmissibility of 1.5-1.7 times that of primary cases infected with other lineages. The increased transmissibility of B.1.1.7 was multiplicative across age and viral load.

**Conclusions:** The results found in this study add new knowledge that can be used to mitigate the further spread of SARS-CoV-2 lineage B.1.1.7, which is becoming increasingly widespread in numerous countries. Our results clarify that the transmissibility of B.1.1.7 should be included as a multiplicative effect in mathematical models used as a tool for decision makers. The results may have important public health implications, as household transmission may serve as a bridge between otherwise separate transmission domains, such as schools and physical workplaces, despite implemented non-pharmaceutical interventions.

## 2 Introduction

Control of the current pandemic caused by Severe Acute Respiratory Syndrome Coronavirus 2 (SARS-CoV-2) is increasingly challenged by the emerging variants of concern (VOC). These include lineages associated with increased transmissibility (du Plessis et al., 2021; Tang et al., 2020; Volz et al., 2021), severe outcomes such as hospitalization (NERV-TAG, 2021; Bager et al., 2021), and/or mortality (Challen et al., 2021; Davies et al., 2021b) and/or whether they can escape immune protection by natural immunization (Chen et al., 2021). Variants, such as SARS-CoV-2 VOC 202012/01 (also known as clade 20I/501Y.V1 or lineage B.1.1.7), are identified by whole genome sequencing (WGS) (Volz et al., 2021). The B.1.1.7 lineage was first identified in the southeast of England in September 2020 (Volz et al., 2021). Since then, it has spread quickly to other countries, and is now a dominant strain in large parts of the world (Alpert et al., 2021; Gozzi et al., 2021). In Denmark, B.1.1.7 was first detected on November 14, 2020, and by March 2021 comprised more than 90% of the circulating lineages (Danish Covid-19 Genome Consortium, 2021). As a consequence of the increased transmissibility of lineage B.1.1.7, nonpharmaceutical interventions (NPIs), such as physical distancing and other restrictions, have been shown to be less effective for sustaining epidemic control (Di Domenico et al., 2021).

Increased transmissibility of B.1.1.7 was estimated in models that use data from community-based surveillance with limited metadata. The estimated increased transmissibility of B.1.1.7 range from 35% to 130% across countries (Davies et al., 2021a; Leung et al., 2021; Washington et al., 2021; Zhao et al., 2021). In Denmark, it was estimated to be 36%-55% higher than other circulating lineages (SSI, 2021a,b). These estimates are sensitive to country-specific conditions, such as other circulating lineages, implemented NPIs, and contact tracing efforts, which can all affect the generation time.

Most studies of B.1.1.7 transmission have not addressed transmission in specific settings, e.g., households, and have not included detailed explanatory variables known to affect transmissibility, such as age of primary cases, age of exposed individuals, and viral load of primary case.

Household members live close together and typically share kitchen, bathroom, and common rooms. Thus, close contact is difficult to limit within households, and may present a challenge for epidemic control. Therefore, studies of transmission in the household domain serve as an opportunity to learn about transmission patterns. Furthermore, household transmission may serve as a bridge between otherwise separate transmission domains, such as schools and physical workplaces, despite implemented NPIs.

Denmark has one of the highest SARS-Cov-2 real-time reverse transcription polymerase chain reaction (RT-PCR) testing and WGS capacities in the world. Furthermore, tests for SARS-CoV-2 are free of charge and testing is widespread with current levels of testing exceeding 30,000 weekly tests per 100,000 persons. Moreover, there is comprehensive social insurance, and SARS-CoV-2 sick leave is fully reimbursed. Hence, neither access to tests nor financial reasons represent major barriers to obtaining a test. Since December 2020, it has been a government policy to use WGS data for surveillance of the Danish epidemic. This has resulted in more than 70% of all RT-PCR positive tests being selected for WGS since January 11, 2021.

The aim of this study was to estimate the household transmissibility SARS-CoV-2 for lineage B.1.1.7 compared with other lineages, by age and viral load. Furthermore, we wanted to estimate whether there is a multiplicative or additive effect of the increased transmissibility of B.1.1.7 compared with other lineages.

## 3 Data and Methods

### 3.1 Register Data

We used comprehensive Danish register data, comprising the full population of Denmark, all RT-PCR tests for SARS-CoV-2 from the Danish Microbiology Database (MiBa), and all positive RT-PCR tests that were sampled for WGS. We used the Danish civil registration number, which is a unique personal identifier, to link positive and negative RT-PCR tests to a national registry of address codes. Thereby, we established a data set of all Danish households, which enabled analysis of presumed household transmission by age, Ct value and SARS-CoV-2 lineage. For a further description of this procedure, see Lyngse et al. (2020).

In Appendix A, we provide descriptive statistics from December 20, 2020 (week 52) to February 21, 2021 (week 7) to provide background information for our choice of study sample.

#### 3.1.1 Study Data

We restricted our study sample to comprise primary cases identified in the study period from January 11 (week 2) to February 7, 2021 (week 5). We allowed for 14 days follow up for secondary cases to occur. There were no changes in public health measures or COVID-19 related restrictions in this period, and the period did not include any public holidays. Week 52 (2020) and week 1 (2021) were affected by Christmas and New Year, while schools opened for grades 0-4 (ages 6-10 years) in week 6. We further restricted our study sample to households with two to six members in order to have relatively comparable households, and thus we excluded, e.g., long term care facilities and other residential institutions.

### 3.2 Whole Genome Sequencing (WGS)

During the study period, RT-PCR tests for SARS-CoV-2 could be obtained from either community testing facilities at TestCenter Denmark (TCDK) or in hospitals, which serve patients and healthcare personnel. All samples from TCDK were analyzed at Statens Serum Institut (SSI), whereas samples from hospitals were analyzed at the hospitals’ departments of clinical microbiology. Testing through TCDK accounted for approximately 75% of all tests and 70% of all positive tests in Denmark (Lyngse et al., 2021). Furthermore, TCDK has used the same protocol for RT-PCR across the full study period. Sequencing of the genome of SARS-CoV-2 was carried out by The Danish COVID-19 Genome Consortium, which was established in March 2020 with the purpose of assisting public health authorities by providing rapid genomic monitoring of the spread of SARS-CoV-2.

As not all positive samples have been selected for WGS, it is important to understand the sample selection process. Information on WGS sample selection criteria and Ct values was only available for positive cases that were identified through TCDK. On January 11, 2021 (week 2), SSI started systematic selection of positive samples for WGS using a Ct value cut-off, in order to maximize the probability of a suitable genome for WGS analysis. During week 2, SSI used a cut-off of Ct<30, Ct<32, and Ct<35. In week 3-6, SSI used a cut-off of Ct<35. During periods with excess WGS capacity, SSI included samples with higher Ct values (35<Ct≤38). An RT-PCR test is positive, if Ct≤38. This is supported by the data (Figure S3 and S4).

#### 3.2.1 Sample selection bias

In our data, not all positive cases have a successfully sequenced genome. This can be due to various reasons, e.g., sequencing capacity constraints. Moreover, the probability of successfully sequencing a genome is correlated with the viral load, which is reflected in the Ct value. Therefore, sample selection bias is a major concern. If some cases have a higher probability of being selected for WGS than others, it can lead to false conclusions. In Appendix A, we provide summary statistics to substantiate our choice of study period. As both viral load (Ct values) and age of the primary case are associated with transmissibility (Lyngse et al., 2021; Lee et al., 2021; Marks et al., 2021), we naturally explored this.

### 3.3 Statistical Analyses

We defined primary cases as the first identified RT-PCR positive SARS-CoV-2 case in a household, and any cases that were detected in the same household within the following 1-14 days were considered to be secondary cases (see also sensitivity analysis of this below). If more than one person tested positive on the first date, the primary case was randomly selected. We utilized two concepts for transmissibility of the primary case: transmission risk and transmission rate. The *transmission risk* describes the risk of infecting at least one other person within the household, and equals one if any (one or more) secondary cases are identified within the same household, and zero otherwise. The *transmission rate* is the proportion of potential secondary cases within the same household that tested positive. The two transmissibility measures are weighted on the primary case level, such that each primary has a weight of one.

Furthermore, we utilized one concept for susceptibility of the potential secondary case: attack rate. The (secondary) attack rate is defined as the proportion of potential secondary cases that tested positive. The attack rate is weighted on the potential secondary case level, such that each potential secondary case has a weight of one.

We estimated the transmission rate and transmission risk for each 10 year age group separately and stratified by lineage B.1.1.7 and other lineages.

To investigate whether the increased transmissibility of B.1.1.7 compared with other lineages was best described as an additive or multiplicative effect, we compared the model fit of both a linear and a logistic regression analysis, using the Akaike Information Criteria (AIC).

We used a logistic regression model to estimate the odds ratio of the transmission rate and transmission risk for B.1.1.7 compared with other lineages. As the transmissibility can be dependent on the age of the primary case, the age of the potential secondary case, and the viral load (measured by cycle threshold (Ct) value) (Lyngse et al., 2021; Lee et al., 2021; Marks et al., 2021) were included as explanatory variables.

See Appendix C for further details of the statistical analyses.

#### 3.3.1 Sensitivity Analyses

To investigate the robustness of the estimated transmissibility across age groups, we supplemented our main analyses of ten-year age groups with five-year age groups.

We estimated the transmission rate and transmission risk by Ct value intervals.

The estimates are sensitive to the definition of primary and secondary cases. In our approach, it is possible that a co-primary case may be misclassified as a secondary case, if she is tested positive one or more days later than the first identified case. In order to investigate the robustness of the results to the definition of primary and secondary cases, we additionally analyzed the data defining secondary cases as those that tested positive at 1-14 days (as in the main analysis), 2-14 days, 3-14 days and 4-14 days after the primary case.

### 3.4 Ethical statement

This study was conducted on administrative register data. According to Danish law, ethics approval is not needed for such research. All data management and analyses were carried out on the Danish Health Data Authority’s restricted research servers with project number FSEID-00004942. The publication only contains aggregated results and no personal data. The publication is, therefore, not covered by the European General Data Protection Regulation.

## 4 Results

Within the study period, a total of 8,093 household primary cases were identified, of which 82% (6,632) were selected for WGS, and 65% (5,241) generated a high-quality SARS-CoV-2 genome (Table 1). Lineage B.1.1.7 was found in 15% (808) of these genomes. The primary cases lived in households comprising 2-6 persons with a total of 16,612 potential secondary cases, of which 4,133 tested positive. This implies an attack rate of 25% (4,133/16,612).

**Table 1:** Summary Statistics

|  | Primary Cases |  |  |  | Potential | Positive | Attack |
| --- | --- | --- | --- | --- | --- | --- | --- |
|  | Total | Selected<br>for WGS | With<br>Genome | With<br>B.1.1.7 | Secondary<br>Cases | Secondary<br>Cases | Rate<br>(%) |
| **Total** | 8,093 | 6,632 | 5,241 | 808 | 16,612 | 4,133 | 25 |
| **Sex** |  |  |  |  |  |  |  |
| Male | 3,648 | 3,013 | 2,406 | 419 | 8,905 | 2,190 | 25 |
| Female | 4,445 | 3,619 | 2,835 | 389 | 7,707 | 1,943 | 25 |
| **Age** |  |  |  |  |  |  |  |
| 0-10 | 419 | 327 | 237 | 54 | 3,490 | 822 | 24 |
| 10-20 | 795 | 670 | 557 | 91 | 3,270 | 755 | 23 |
| 20-30 | 1,531 | 1,294 | 1,020 | 204 | 2,347 | 494 | 21 |
| 30-40 | 1,353 | 1,101 | 870 | 143 | 1,876 | 483 | 26 |
| 40-50 | 1,464 | 1,182 | 920 | 119 | 2,167 | 521 | 24 |
| 50-60 | 1,443 | 1,166 | 917 | 132 | 2,020 | 536 | 27 |
| 60-70 | 669 | 539 | 449 | 38 | 919 | 315 | 34 |
| 70-80 | 300 | 255 | 190 | 23 | 392 | 150 | 38 |
| >80 | 119 | 98 | 81 | <5 | 131 | 57 | 44 |
| **Household**<br>**Size** |  |  |  |  |  |  |  |
| 2 | 3,308 | 2,716 | 2,108 | 298 | 3,308 | 1,019 | 31 |
| 3 | 1,886 | 1,549 | 1,235 | 189 | 3,635 | 843 | 23 |
| 4 | 1,848 | 1,486 | 1,178 | 193 | 5,368 | 1,292 | 24 |
| 5 | 790 | 661 | 534 | 92 | 3,042 | 714 | 23 |
| 6 | 261 | 220 | 186 | 36 | 1,259 | 265 | 21 |

Notes: This table provides summary statistics for the number of primary cases, potential secondary cases, positive secondary cases, and attack rates in the study, stratified by sex, age and household sizes. Summary statistics for five-year age groups are shown in Table S2.

The intra-household correlation of lineages between primary and positive secondary cases was investigated using the proportion of positive secondary cases that were infected with the same lineage (B.1.1.7 vs. other lineages) as the primary case (Table 2). For primary cases infected with B.1.1.7, 96% of the positive secondary cases (that were successfully sequenced) were also infected with B.1.1.7. Similarly, for primary cases infected with other lineages, 99% of the positive secondary cases (that were successfully sequenced) were also infected with other lineages. For the primary cases without a successfully sequenced genome, 20% of the positive secondary cases (that were successfully sequenced) were infected with B.1.1.7 and 80% with other lineages. This distribution roughly corresponds to the underlying prevalence in the community during period of the study.

**Table 2:** Intra-household correlation of lineages between primary and positive secondary cases

| — Primary cases — |  | — Positive secondary cases — |  |  |  | Potential | Attack |
| --- | --- | --- | --- | --- | --- | --- | --- |
| Lineage | N | B.1.1.7 | Other lineages | No Genome | Total | secondary cases | rate (%) |
| B.1.1.7 | 808 | 472 | 19 | 165 | 656 | 1,719 | 38 |
| Other lineages | 4,433 | 18 | 1,750 | 721 | 2,489 | 9,115 | 27 |
| No Genome | 2,852 | 133 | 540 | 315 | 988 | 5,778 | 17 |
| Total | 8,093 | 623 | 2,309 | 1,201 | 4,133 | 16,612 | 25 |

Notes: There were 8,093 primary cases, of which 808 (10%) were infected with B.1.1.7, 4,433 (55%) were infected with other lineages, and 2,852 (35%) did not have a successfully sequenced genome. The 808 primary cases infected with B.1.1.7 had 656 positive secondary cases. Of these cases, 75% (472+19=491) were successfully sequenced. Of these, 96% (472) were infected with B.1.1.7 and 4% (19) with other lineages.

In households where the primary cases were infected with B.1.1.7, the attack rate was 38%, compared with 27% when the primary cases were infected with other lineages, and 17% when the primary case did not have a successfully sequenced genome.

The age specific transmissibility followed a U shaped pattern with the lowest transmission from primary cases in the 10 to 30 years age range, higher from younger children, and highest from elderly cases (Figure 1). Both the transmission rate (Figure 1, panel a) and the transmission risk (Figure 1, panel b) were higher for B.1.1.7 (red) compared with other lineages (blue) across all ten-year age groups. The transmissibility was lower for primary cases without a successfully sequenced genome (gray).

**Figure 1:**
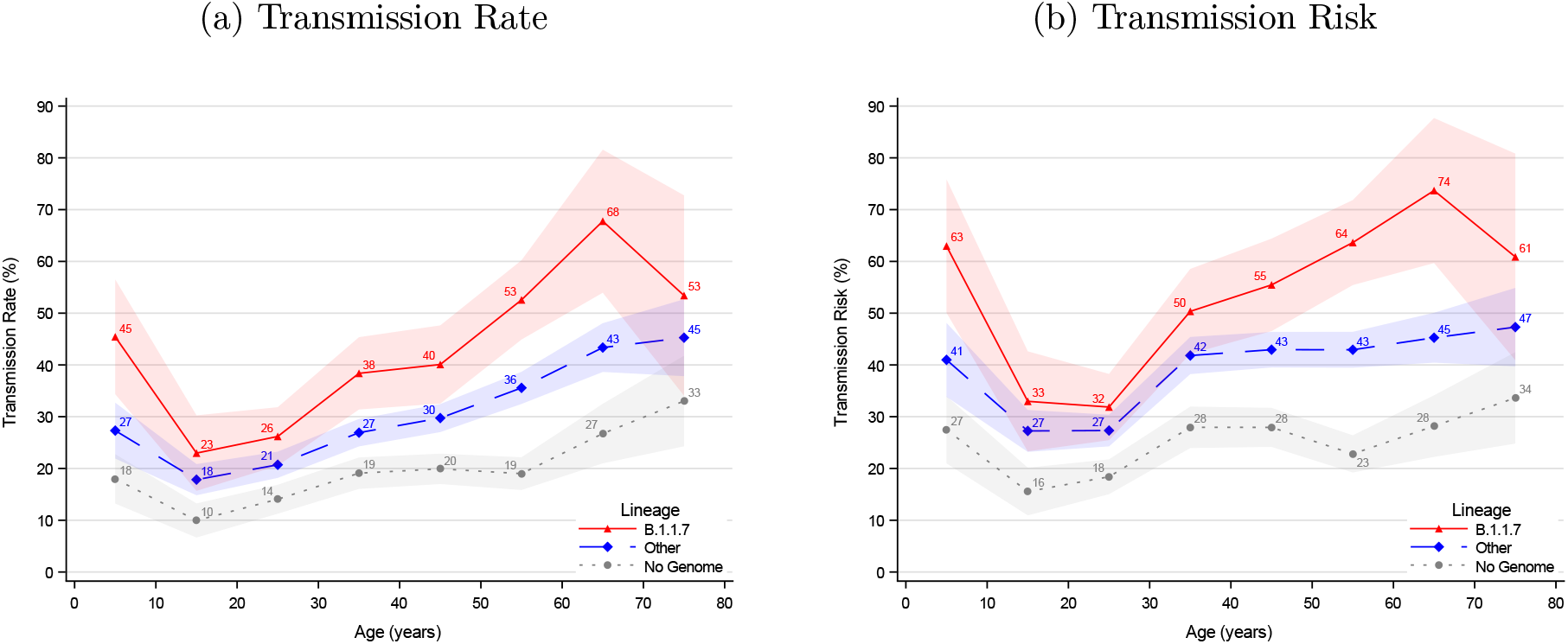
Age structured transmissibility stratified by lineage of the primary case Notes: The transmission rate describes the proportion of potential secondary cases within the household that were infected. The transmission risk describes the proportion of infected primary cases that infected at least one secondary case. Figure S7 provides the same graphs for five-year age groups. The shaded areas show the 95% confidence bands clustered on the household level.

To investigate whether the increased transmissibility of B.1.1.7 compared with other lineages was best described as an additive or multiplicative effect, we compared the model fit of both a linear and a logistic regression analysis. We compared the fit of the two models using the Akaike Information Criteria (AIC) and found that the logit model was a better fit (Appendix C). This supports the hypothesis that the effect of the increased transmissibility is best described as a multiplicative effect.

Using a logit model, we estimated the increased transmission rate and transmission risk for B.1.1.7 compared with other lineages. In Table 3, we present the crude estimates as well as models controlling for age of the primary case, age of the potential secondary cases, and Ct value of the primary case. Primary cases infected with B.1.1.7 were 1.5 times more transmissible than primary cases infected with other lineages, without any adjustments. When controlling for age and viral load, this effect was 1.6.

**Table 3:** Odds ratio estimates for transmissibility for B.1.1.7 compared with other lineages

|  | Transmission Rate |  |  |  | Transmission Risk |  |  |
| --- | --- | --- | --- | --- | --- | --- | --- |
|  | I | II | III | IV | V | VI | VII |
| B.1.1.7 | 1.50 | 1.68 | 1.69 | 1.63 | 1.52 | 1.66 | 1.61 |
| 95%-CI | (1.30-1.72) | (1.46-1.94) | (1.47-1.95) | (1.39-1.91) | (1.31-1.77) | (1.42-1.93) | (1.36-1.90) |
| Constant | ✓ | ✓ | ✓ | ✓ | ✓ | ✓ | ✓ |
| Age, Primary Case |  | ✓ | ✓ | ✓ |  | ✓ | ✓ |
| Age, Pot. Sec. Case |  |  | ✓ | ✓ |  |  |  |
| Ct Value |  |  |  | ✓ |  |  | ✓ |
| Observations | 10,834 | 10,834 | 10,834 | 8,762 | 10,834 | 10,834 | 8,762 |
| Households | 5,241 | 5,241 | 5,241 | 4,172 | 5,241 | 5,241 | 4,172 |

Notes: Columns I-IV provide odds ratio estimates for the increased transmission rate of B.1.1.7 compared with other lineages. Columns V-VII show the same for the transmission risk. Column I provides the crude estimates, i.e., only with a constant and without any controls. Column II further includes fixed effects for ten-year age groups of the primary cases. Column III further includes the age of potential secondary cases. Column IV further includes fixed effects for Ct values in bi-value groups. This sample is further restricted to only include primary cases identified in TCDK, as we only have Ct values on those. Column V provides the crude estimates, i.e., only with a constant and without any controls. Column VI further includes fixed effects for ten-year age groups of the primary cases. Column VII further includes fixed effects for Ct values in bi-value groups. This sample is further restricted to only include primary cases identified in TCDK, as we only have Ct values on those. All effects are included as fixed effects. Pot. Sec. Case = Potential Secondary Cases. Only primary cases identified in TCDK are included in models with Ct values. 95% confidence bands clustered on the household level.

## 5 Discussion and Conclusion

We used national population data to estimate the household transmissibility of the SARS-CoV-2 lineage B.1.1.7 compared with other lineages. We utilized detailed administrative register data comprising the full Danish population and the ability to link data across registers on a person level. This combined with a large proportion of the population being tested, a large national WGS capacity, and an understanding of the sampling selection process, allowed us to estimate the household transmissibility controlling for age and viral load.

We found that B.1.1.7 had a household transmissibility 1.5-1.7 times higher compared with other lineages, which is in line with B.1.1.7 transmissibility estimates from modelling studies of surveillance data (NERVTAG, 2021; Davies et al., 2021b; Piantham & Ito, 2021; Volz et al., 2021).

Furthermore, we estimated the transmissibility across age groups and found that lineage B.1.1.7 generally follows the pattern of other lineages, where teenagers are the least transmissible within households. However, B.1.1.7 was consistently more transmissible per age group compared with other lineages.

We found that the increased transmissibility of B.1.1.7 is a multiplicative effect of the transmissibility of other lineages, rather than an additive effect. Only one previous study has estimated both the additive effect and the multiplicative effect (Graham et al., 2021), but they did not test the two models against each other. The multiplicative effect implies that the known risk factors for increased transmissibility are amplified by 1.5-1.7 times when the case is infected with B.1.1.7.

We have previously found that younger children are more transmissible within the household compared with teenagers (Lyngse et al., 2020, 2021). There is still disagreement about the effect of B.1.1.7 on the transmissibility in children (Rasmussen, 2021; Walker et al., 2021). We here found that children (<10 years)—like adults—also exhibit a higher transmissibility within households if they are infected with B.1.1.7.

The increased transmissibility of 1.5-1.7 times for B.1.1.7 may have public health implications. For example, for contact tracing, this means that cases with a high predicted transmissibility, e.g., by viral load or age (Lyngse et al., 2021; Lee et al., 2021; Marks et al., 2021), that are infected with B.1.1.7 are even more transmissible and thus should be prioritized. Naturally, household contacts are different from other contacts. They are more frequent, closer and of a longer time duration, compared with other settings, such as workplaces. Additionally, many people live with a partner around their own age and parents live with their children. The results underline the importance of timely and efficient management and isolation of confirmed cases to limit transmission in the household domain. Transmission in households may serve as a bridge between otherwise separate domains, such as schools and physical workplaces, despite implemented NPIs in these domains. Moreover, it might be more challenging for young children to maintain social distancing and to adhere to NPIs in general, more outbreaks of B.1.1.7 in kindergardens and primary schools could be expected. This is important for decision makers when making decision about lockdowns and re-openings of parts of society. Furthermore, our results imply that the transmissibility of B.1.1.7 should be modelled as a multiplicative effect and not an additive effect. This is pivotal for the validity and accuracy of simulations models of the current pandemic, which are used as tools for decision makers.

The mechanisms behind the increased transmissibility of B.1.1.7 are not fully elucidated. Recently it has been suggested that enhanced binding of the N501Y mutated spike protein may result in increased binding affinity to the human angiotensin-converting enzyme 2 (ACE2) (Luan et al., 2021; Zhang et al., 2021). Furthermore, Kissler et al. (2021) and Calistri et al. (2021) found that the infectious period for cases infected with B.1.1.7 was generally longer compared with cases infected with other lineages. For children, this has also previously been described for seasonal influenza by Ng et al. (2016). The longer infectious period could contribute to the increased transmissibility of lineage B.1.1.7 There are several strengths in the present study.

This nationwide study is based on detailed administrative data that enabled us to control for individual specific characteristics of both primary and potential secondary cases. Furthermore, we restricted our sample to only include households with 2-6 members during a period with no national holidays, no changes in government restrictions, and systematic sampling for WGS. Furthermore, we challenged our approach by investigating the intra-household correlation of lineages between primary and positive secondary cases. We found that the vast majority of secondary cases were infected with the same lineage (B.1.1.7 vs other lineages) as the primary case. When investigating the intra-household correlation of lineages between primary and positive secondary cases, we found that 96% of the secondary cases associated with a primary case infected with B.1.1.7 were also infected with B.1.1.7. Similarly, we found that 1% of the secondary cases associated with a primary case infected with other lineages were infected with B.1.1.7. This suggests that only a minor fraction of the positive secondary cases were misclassified.

We estimated the increased transmissibility of B.1.1.7 relative to a baseline of other circulating lineages. It is evident that these estimates depend on the composition of this baseline. In our study period, 82% of all positive cases were selected for WGS and 65% of the total cases had a successfully sequenced genome (Tabel S1). The commonly circulating lineages included B.1.258.11, B.1.258, B.1.221.3, B.1.221. B.1.160 and B.1.177 (Figure S1). Therefore, it is not likely that our findings are an artefact generated by a misleading baseline of other lineages.

There is a significant proportion of positive RT-PCR positive samples without a successfully sequenced genome that could not be assigned to specific lineages. This can potentially result in sample selection bias. Samples with low viral load (high Ct values) were less likely to be selected for WGS and successfully sequenced (Figure S3 and S4). Cases with low viral load have been shown to be less transmissible (Lyngse et al., 2021; Lee et al., 2021; Marks et al., 2021). If cases infected with B.1.1.7 have higher viral loads than cases infected with other lineages, this would lead to over-sampling of cases infected with B.1.1.7. Presently, this is not fully elucidated. Calistri et al. (2021) have found that cases infected with B.1.1.7 have a higher viral load, whereas Kissler et al. (2021) and the present study (Figure S5) found no difference. This implies that over-sampling of cases infected with B.1.1.7 was not a problem in this study. Furthermore, we controlled for Ct values in our multivariable regression model, and this confirmed that B.1.1.7 was associated with increased transmission even after adjusting for Ct values.

There were only relatively minor changes in the estimates of the increased transmissibility of B.1.1.7 compared with other lineages when varying the controls. This suggests that the increased transmissibility of B.1.1.7 is independent of the age of the infected person, age of the exposed person and Ct value. Moreover, the estimates could be sensitive to the definition of primary and secondary cases. However, when we restricted our analysis to only include secondary cases identified on days 1-14, 2-14, 3-14, and 4-14, we found no significant changes in the estimates. This demonstrates that the estimates of the increased transmissibility of B.1.1.7 were not dependent on the inclusion criteria for secondary cases.

Some limitations apply to this study. This is a retrospective observational study, therefore causality naturally cannot be inferred. Additionally, we did not have access to data on rapid antigen tests, which have been increasingly used in Denmark since December 2020. All cases with a positive antigen test were recommended to have a confirmatory RT-PCR test. If cases tested positive with an antigen test and not a RT-PCR test, we could not include these as positive cases. Despite of these limitations, we believe that the results of this study provide useful new insights into the transmissibility of B.1.1.7.

In summary, we found an attack rate of 38% in households with a primary cases infected with B.1.1.7 and 27% in households with a primary case infected with other lineages. Primary cases infected with B.1.1.7 had an increased transmissibility of 1.5-1.7 times that of primary cases infected with other lineages. The increased transmissibility of B.1.1.7 is multiplicative across age and viral load.

The spread of lineage B.1.1.7 has been explosive in countries across the world. The results found in this study add new knowledge that can be used to mitigate the further spread of SARS-CoV-2 lineage B.1.1.7. Further studies are needed to evaluate the transmissibility in other settings, such as workplaces, schools and other places of infection.

## Data Availability

Statens Serum Institut and The Danish Health Data Authority.

## Appendix A: Descriptive Statistics

From December 21, 2020 (week 52) to February 21, 2021 (week 7), Denmark had 68,169 SARS-CoV-2 cases identified with RT-PCR, of which, 35,684 (52%) were selected for WGS and 28,383 (42%) came back with a genome (Table S1).

**Table S1:**
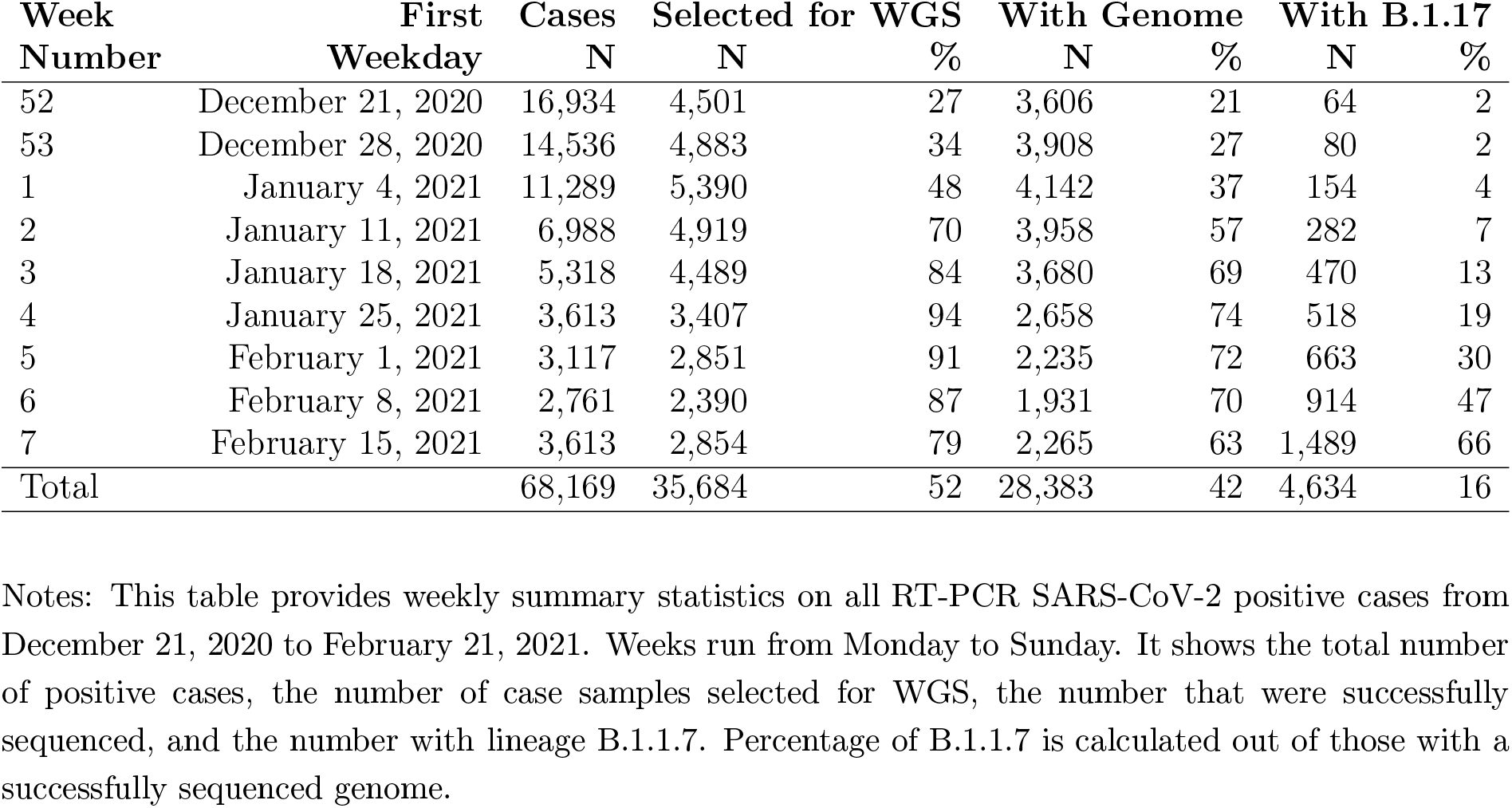
Summary statistics for all positive cases

| Week<br>Number | First<br>Weekday | Cases<br>N | Selected for WGS<br>N | % | With Genome<br>N | % | With B.1.17<br>N | % |
| --- | --- | --- | --- | --- | --- | --- | --- | --- |
| 52 | December 21, 2020 | 16,934 | 4,501 | 27 | 3,606 | 21 | 64 | 2 |
| 53 | December 28, 2020 | 14,536 | 4,883 | 34 | 3,908 | 27 | 80 | 2 |
| 1 | January 4, 2021 | 11,289 | 5,390 | 48 | 4,142 | 37 | 154 | 4 |
| 2 | January 11, 2021 | 6,988 | 4,919 | 70 | 3,958 | 57 | 282 | 7 |
| 3 | January 18, 2021 | 5,318 | 4,489 | 84 | 3,680 | 69 | 470 | 13 |
| 4 | January 25, 2021 | 3,613 | 3,407 | 94 | 2,658 | 74 | 518 | 19 |
| 5 | February 1, 2021 | 3,117 | 2,851 | 91 | 2,235 | 72 | 663 | 30 |
| 6 | February 8, 2021 | 2,761 | 2,390 | 87 | 1,931 | 70 | 914 | 47 |
| 7 | February 15, 2021 | 3,613 | 2,854 | 79 | 2,265 | 63 | 1,489 | 66 |
| Total |  | 68,169 | 35,684 | 52 | 28,383 | 42 | 4,634 | 16 |

Notes: This table provides weekly summary statistics on all RT-PCR SARS-CoV-2 positive cases from December 21, 2020 to February 21, 2021. Weeks run from Monday to Sunday. It shows the total number of positive cases, the number of case samples selected for WGS, the number that were successfully sequenced, and the number with lineage B.1.1.7. Percentage of B.1.1.7 is calculated out of those with a successfully sequenced genome.

Lineage B.1.1.7 became increasingly dominant, crowding out other lineages, from December 2020 to February 21, 2021, (Figure S1).

**Figure S1:**
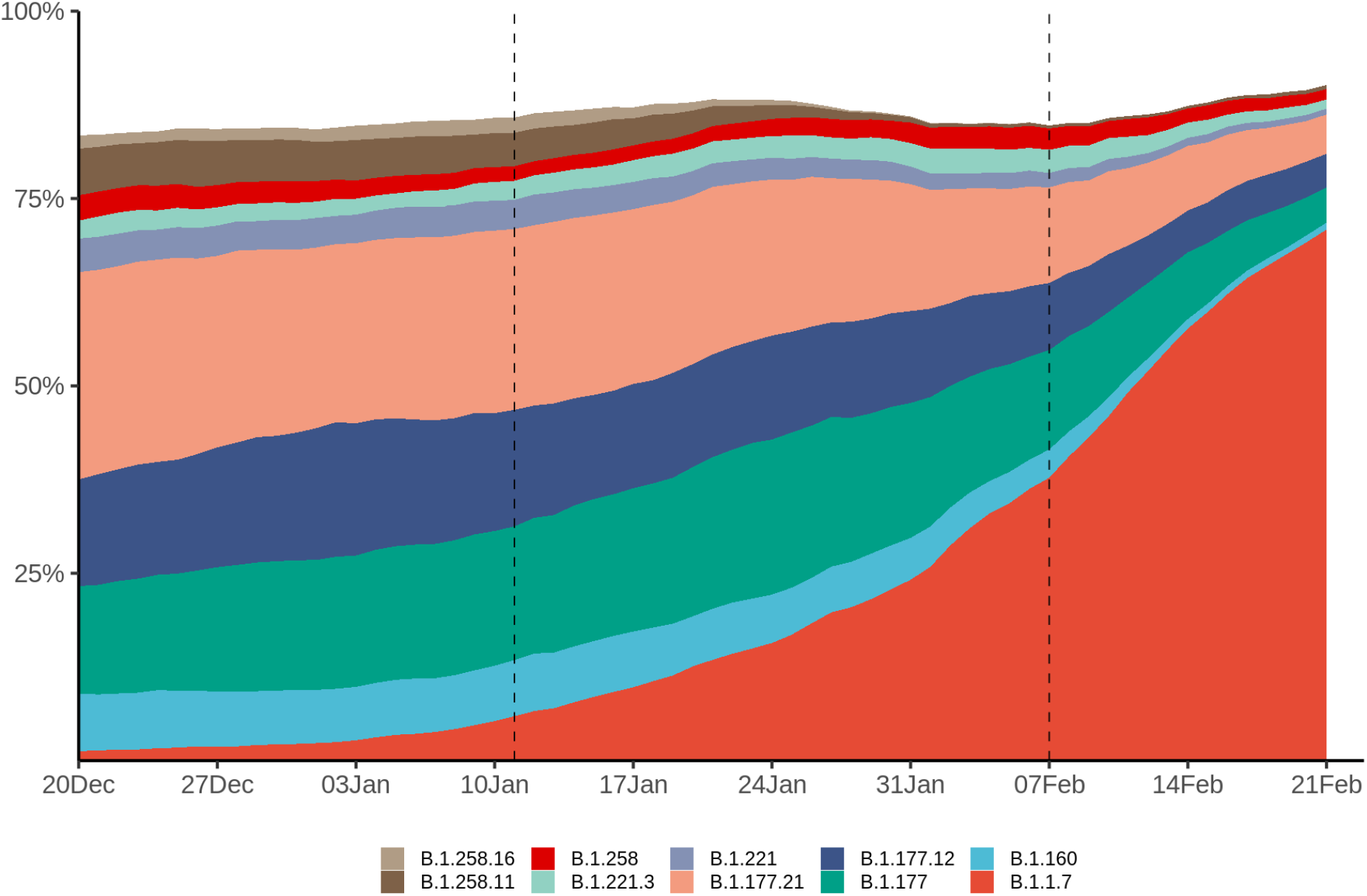
Frequency of detected WGS strains in Denmark over time. Notes: This figure shows the ten most abundant lineages for cases with a complete genome in Denmark during the study period. Less abundant lineages are included in the white space. 14-day rolling average.

The proportion of cases being sampled varied over time depending on whether the cases occurred in TCDK or in hospitals (Figure S2).

**Figure S2:**
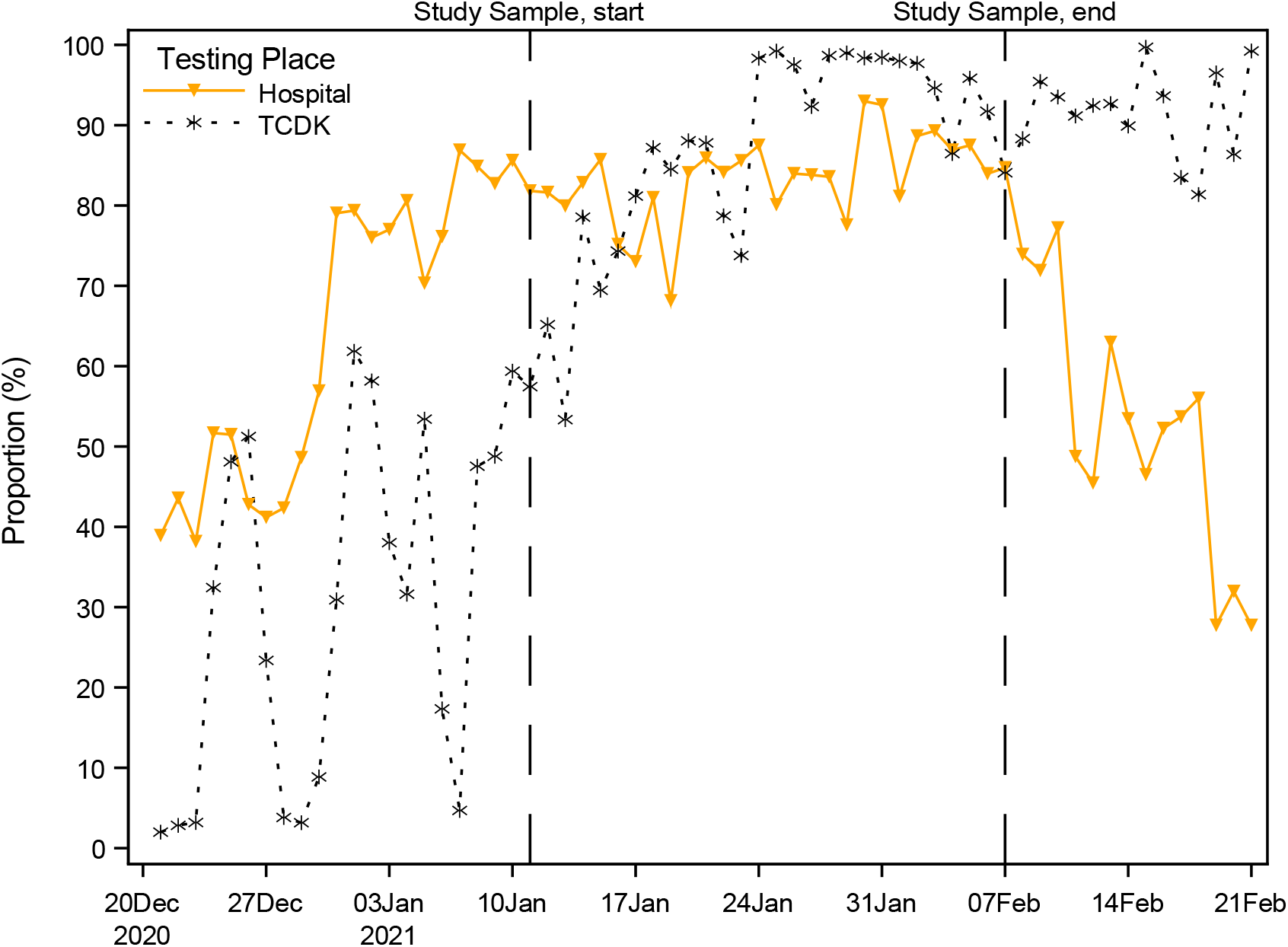
Proportion of positive RT-PCR tests sampled for WGS, stratified by testing facility Notes: This figure shows the proportion of positive RT-PCR test cases that were selected for WGS stratified by testing facility (TCDK or hospital).

The proportion of cases selected for WGS and the proportion that came back with a genome is dependent on the Ct value (Figure S3). For positive tests with a Ct value of 18, 51% of the samples were selected for WGS (purple) and 45% came back with a genome (green). Thus, the success rate was 88% (45/51). Similarly, for positive tests with a Ct value of 38, 26% of the samples were selected for WGS and 5% came back with a genome. Thus, the success rate was 19% (5/26). The success rate starts to decline for tests with a Ct value ≥ 30.

**Figure S3:**
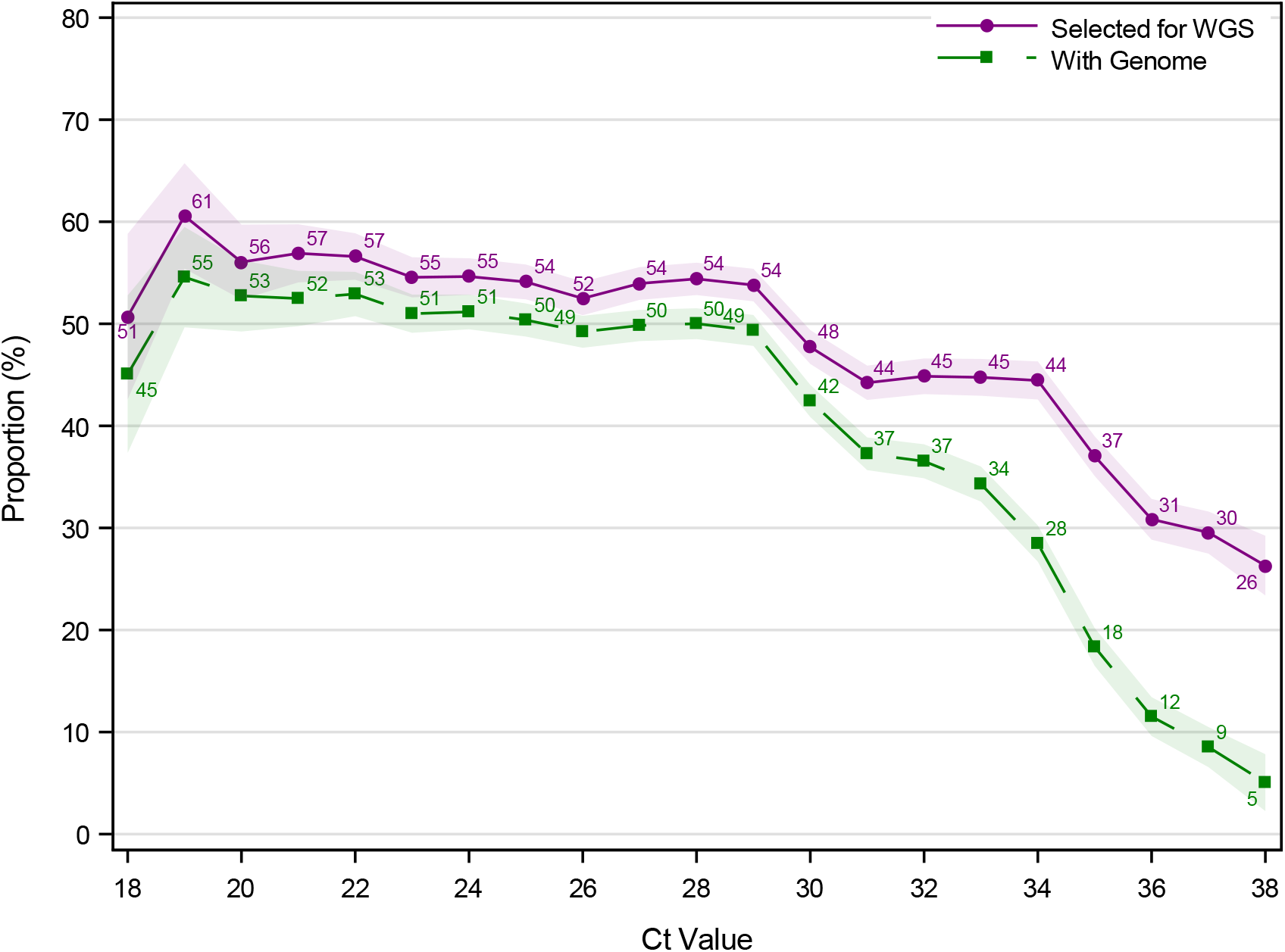
Proportion of positive RT-PCR tests sampled for WGS and with a successfully sequenced genome, by Ct value Notes: This figure shows the proportion of cases selected for WGS and the proportion that were successfully sequenced stratified by the Ct value of the sample. For positive tests with a Ct value of 18, 51% of the samples were selected for WGS (purple) and 45% came back with a genome (green). Thus, the success rate was 88% (45/51). Similarly, for positive tests with a Ct value of 38, 26% of the samples were selected for WGS and 5% came back with a genome. Thus, the success rate was 19% (5/26). Only samples from TCDK are included. An RT-PCR test is positive if the Ct value is ≤38. The shaded areas show the 95% confidence bands.

The proportion of cases being sampled for WGS dependent on the Ct value varies over time (Figure S4). In week 2 TCDK started to sample systematically and to sample on Ct values. From Figure S4, we see that in week 2, TCDK used a Ct value cut-off of 30, 32, and 35. In weeks 3-6, TCDK used a Ct value cut-off of 35. Samples with higher Ct values (35<Ct≤38) were included, when WGS capacity allowed for it.

**Figure S4:**
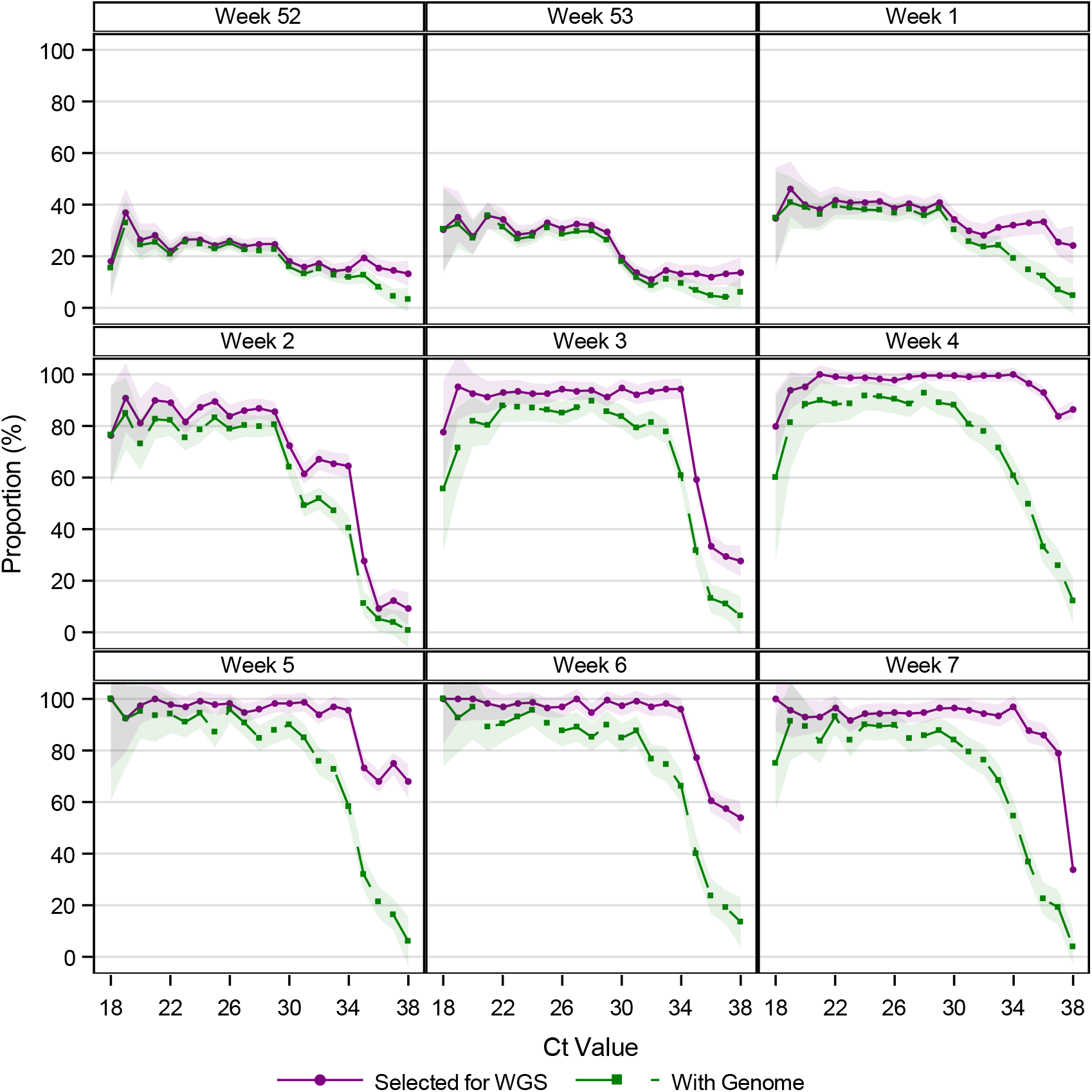
Proportion of positive RT-PCR tests sampled for WGS and with a genome, by Ct value and calendar week Notes: This figures shows the proportion of cases selected for WGS and the proportion that were successfully sequenced stratified by the Ct value of the sample, across weeks. Only samples from TCDK are included. An RT-PCR test is positive if the Ct value is ≤38. The shaded areas show the 95% confidence bands.

The distribution of Ct values of the cases stratified by B.1.1.7 (red), other lineages (blue) are relatively similar, while samples with no genome (gray) have a distribution with higher Ct values (Figure S5).

WGS was mainly obtained for samples with low Ct values compared with the distribution of Ct values from the whole population (gray dashed line in Figure S5). We found that the Ct value distribution for B.1.1.7 and other lineages were approximately similar from week 1 to week 7 (Figure S5). We see a clear shift in the distribution of cases without a successfully sequenced genome from week 2, when SSI started to systematically select case samples on Ct values.

**Figure S5:**
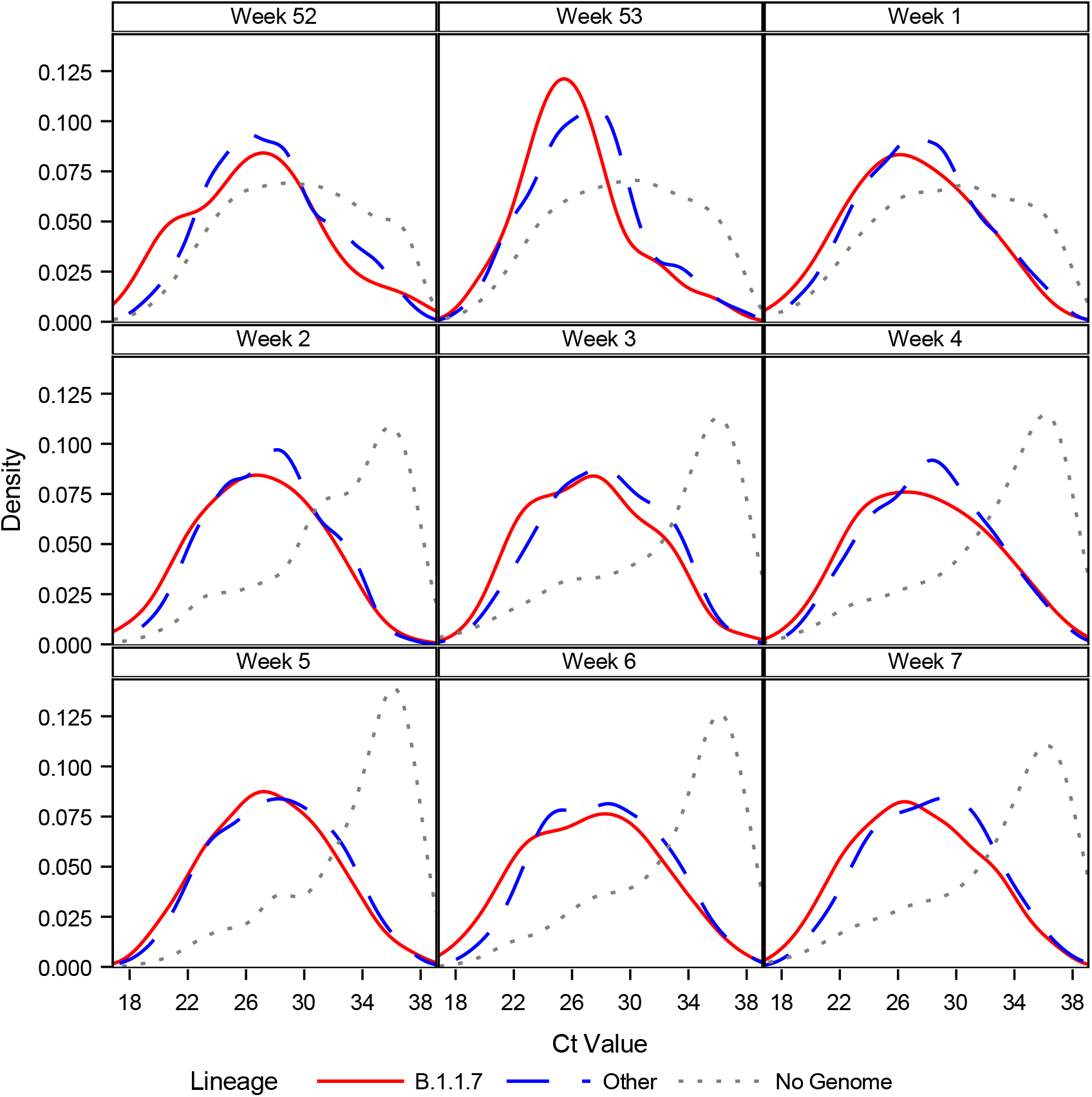
Ct Value distributions by calendar week Notes: This figure shows the kernel density distributions of the Ct value for cases infected with B.1.1.7 (red), other lineages (blue), and without a successfully sequenced genome (gray). In week 2, 2021, SSI started systematic sampling on Ct values from tests from TCDK. Only samples from TCDK are included. An RT-PCR test is positive if the Ct value is ≤38.

The distribution of the age of the cases stratified by B.1.1.7 (red), other lineages (blue) are relatively similar, although B.1.1.7 seems to mainly infect younger people in weeks 2-4 (Figure S6).

**Figure S6:**
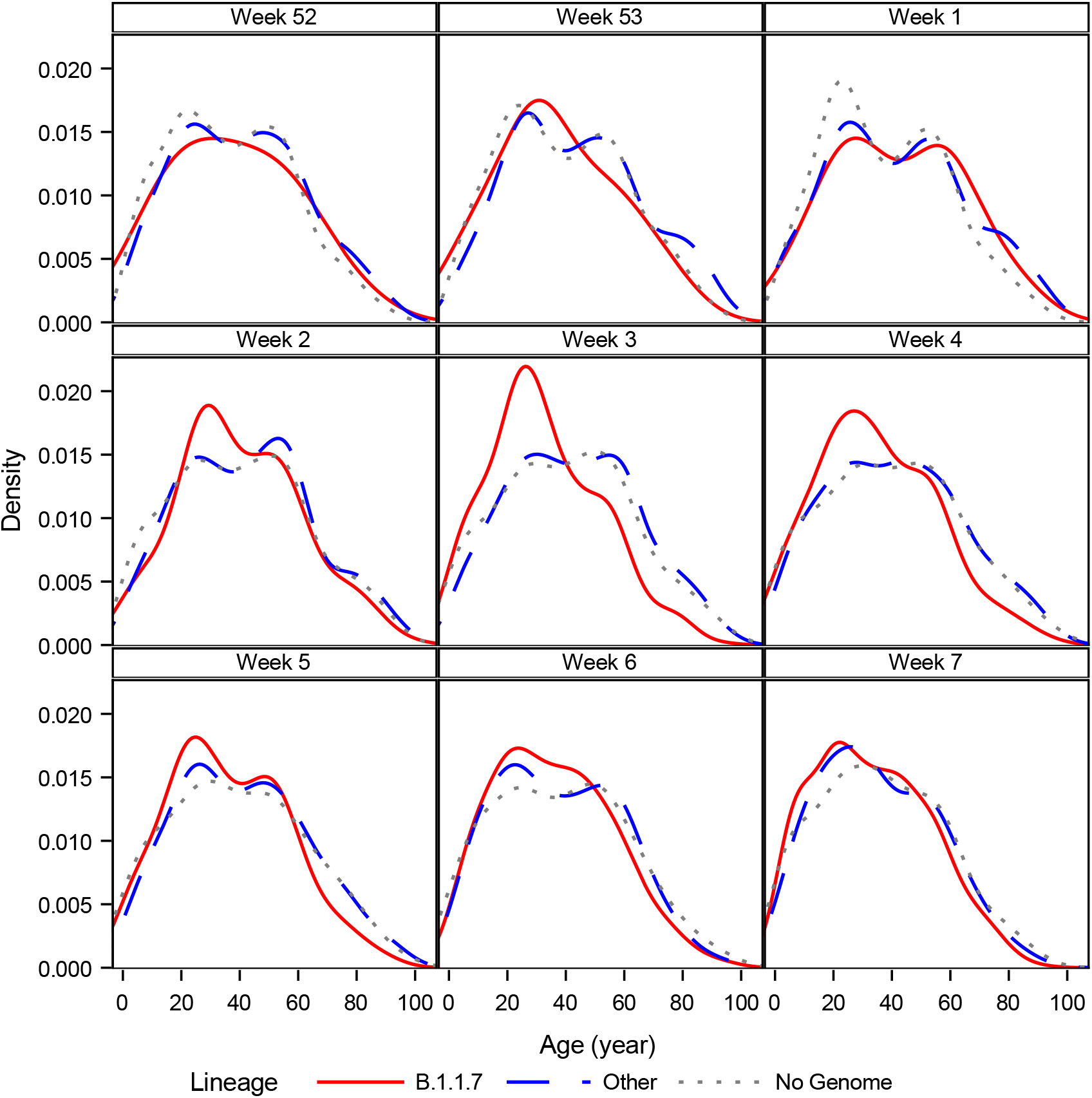
Age distributions by calendar week Notes: This figure shows kernel density distributions of the age for cases infected with B.1.1.7 (red), other lineages (blue), and without a successfully sequenced genome (gray).

## Appendix B: Additional Analyses

**Table S2:**
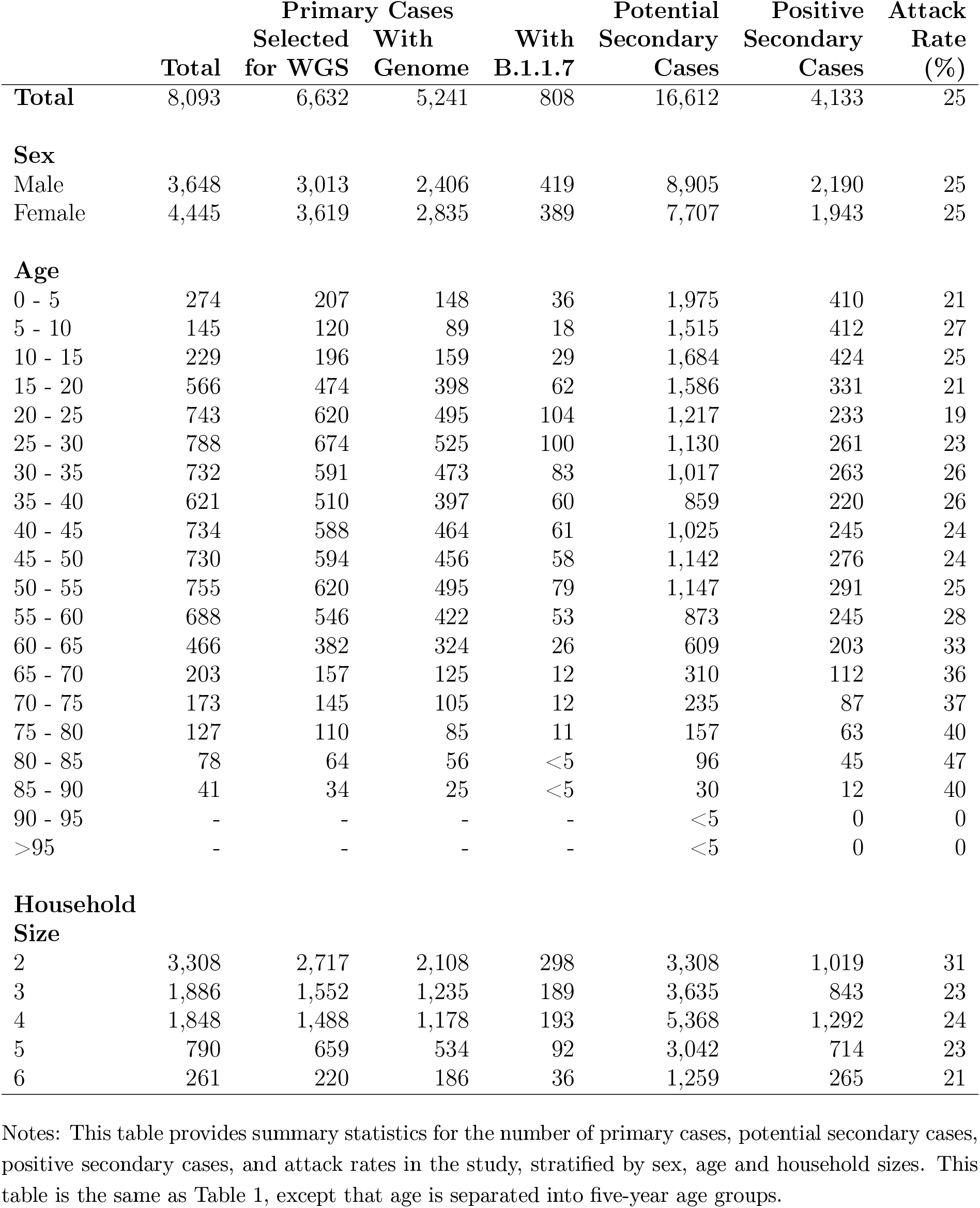
Summary statistics

|  |  | Primary Cases |  |  | Potential | Positive | Attack |
| --- | --- | --- | --- | --- | --- | --- | --- |
|  | Total | Selected<br>for WGS | With<br>Genome | With<br>B.1.1.7 | Secondary<br>Cases | Secondary<br>Cases | Rate<br>(%) |
| **Total** | 8,093 | 6,632 | 5,241 | 808 | 16,612 | 4,133 | 25 |
| **Sex** |  |  |  |  |  |  |  |
| Male | 3,648 | 3,013 | 2,406 | 419 | 8,905 | 2,190 | 25 |
| Female | 4,445 | 3,619 | 2,835 | 389 | 7,707 | 1,943 | 25 |
| **Age** |  |  |  |  |  |  |  |
| 0 - 5 | 274 | 207 | 148 | 36 | 1,975 | 410 | 21 |
| 5 - 10 | 145 | 120 | 89 | 18 | 1,515 | 412 | 27 |
| 10 - 15 | 229 | 196 | 159 | 29 | 1,684 | 424 | 25 |
| 15 - 20 | 566 | 474 | 398 | 62 | 1,586 | 331 | 21 |
| 20 - 25 | 743 | 620 | 495 | 104 | 1,217 | 233 | 19 |
| 25 - 30 | 788 | 674 | 525 | 100 | 1,130 | 261 | 23 |
| 30 - 35 | 732 | 591 | 473 | 83 | 1,017 | 263 | 26 |
| 35 - 40 | 621 | 510 | 397 | 60 | 859 | 220 | 26 |
| 40 - 45 | 734 | 588 | 464 | 61 | 1,025 | 245 | 24 |
| 45 - 50 | 730 | 594 | 456 | 58 | 1,142 | 276 | 24 |
| 50 - 55 | 755 | 620 | 495 | 79 | 1,147 | 291 | 25 |
| 55 - 60 | 688 | 546 | 422 | 53 | 873 | 245 | 28 |
| 60 - 65 | 466 | 382 | 324 | 26 | 609 | 203 | 33 |
| 65 - 70 | 203 | 157 | 125 | 12 | 310 | 112 | 36 |
| 70 - 75 | 173 | 145 | 105 | 12 | 235 | 87 | 37 |
| 75 - 80 | 127 | 110 | 85 | 11 | 157 | 63 | 40 |
| 80 - 85 | 78 | 64 | 56 | <5 | 96 | 45 | 47 |
| 85 - 90 | 41 | 34 | 25 | <5 | 30 | 12 | 40 |
| 90 - 95 | - | - | - | - | <5 | 0 | 0 |
| >95 | - | - | - | - | <5 | 0 | 0 |
| **Household**<br>**Size** |  |  |  |  |  |  |  |
| 2 | 3,308 | 2,717 | 2,108 | 298 | 3,308 | 1,019 | 31 |
| 3 | 1,886 | 1,552 | 1,235 | 189 | 3,635 | 843 | 23 |
| 4 | 1,848 | 1,488 | 1,178 | 193 | 5,368 | 1,292 | 24 |
| 5 | 790 | 659 | 534 | 92 | 3,042 | 714 | 23 |
| 6 | 261 | 220 | 186 | 36 | 1,259 | 265 | 21 |

Notes: This table provides summary statistics for the number of primary cases, potential secondary cases, positive secondary cases, and attack rates in the study, stratified by sex, age and household sizes. This table is the same as Table 1, except that age is separated into five-year age groups.

**Figure S7:**
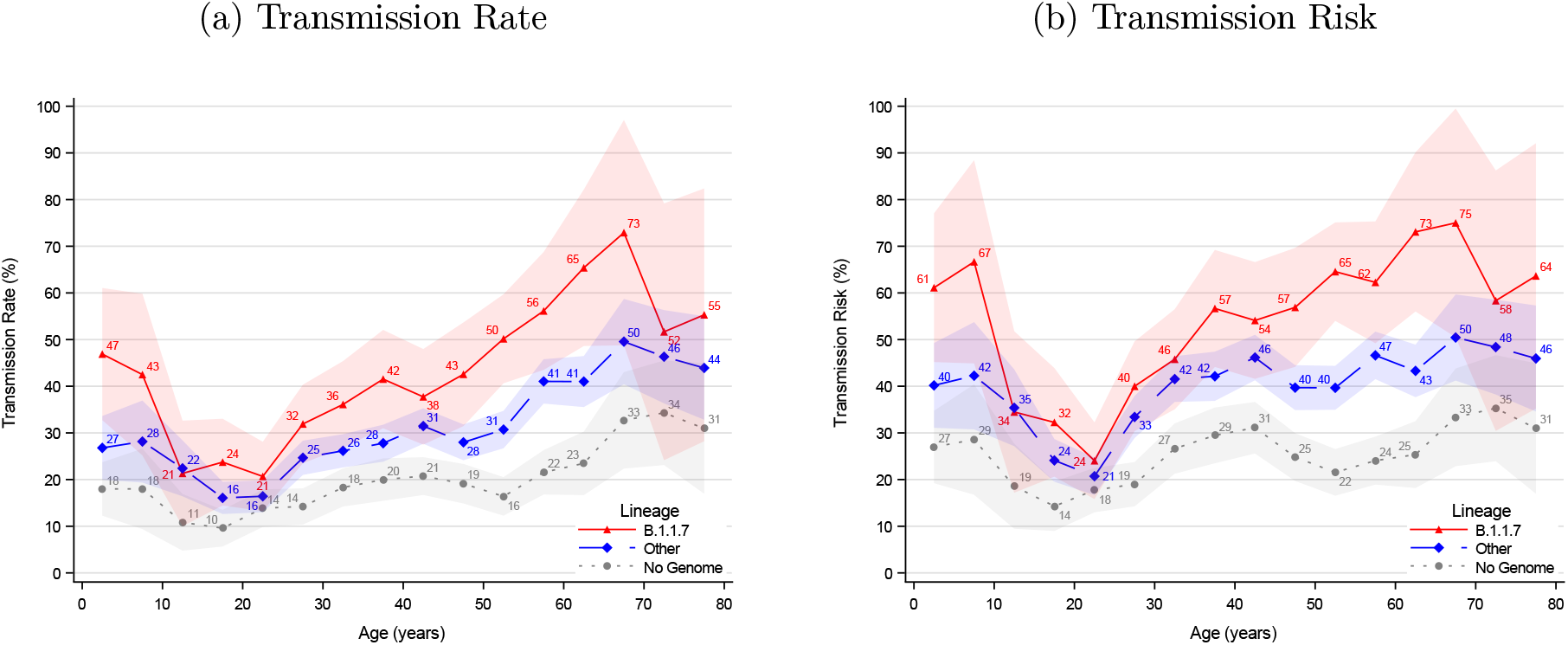
Age structured transmissibility stratified by lineage in five-year age groups. Notes: The transmission rate describes the proportion of potential secondary cases within the household that were infected. The transmission risk describes the proportion of infected primary cases that infected at least one secondary case. This figure is the same as Figure 1, except that it shows five-year age groups. The shaded areas show the 95% confidence bands clustered on the household level.

Primary cases infected with B.1.1.7 generally had a higher transmissibility compared with cases infected with other lineages, across Ct values (Figure S8 and S9).

**Figure S8:**
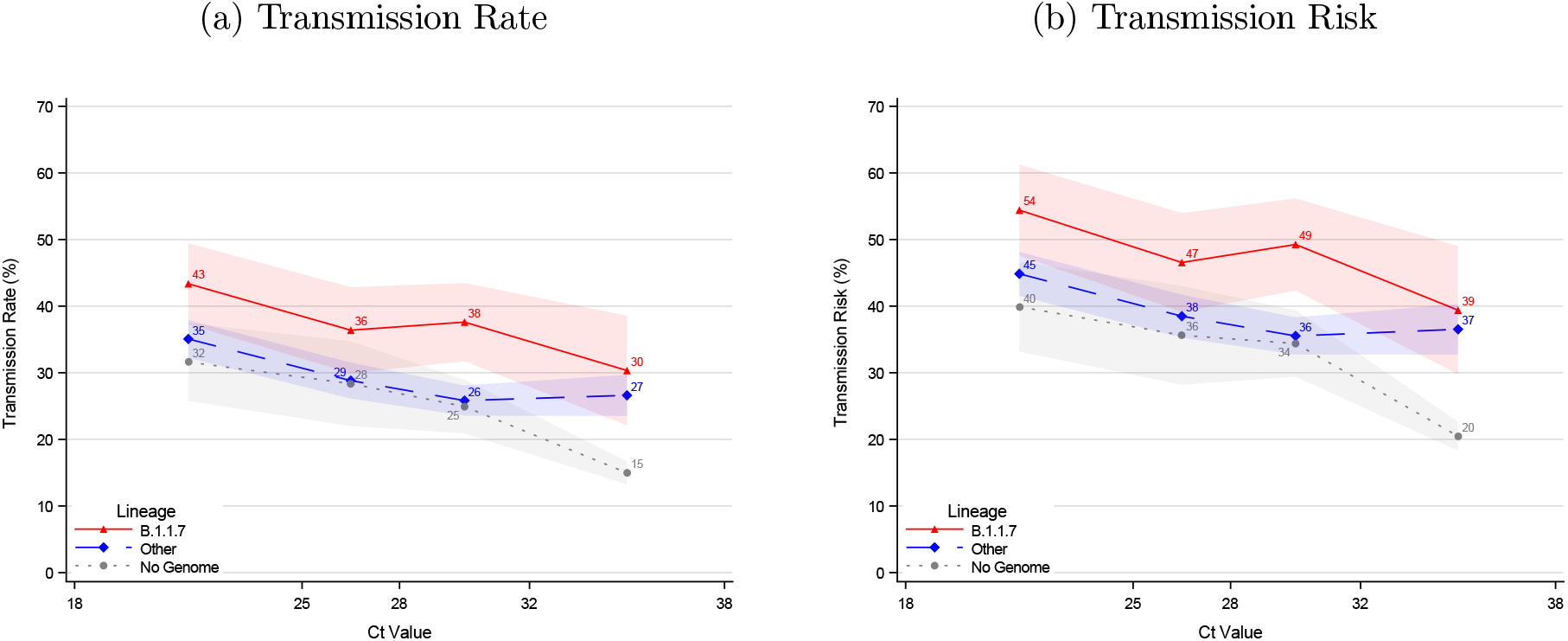
Transmissibility stratified by lineage and Ct value quartiles Notes: The transmission rate describes the proportion of potential secondary cases within the household that were infected. The transmission risk describes the proportion of infected primary cases that infected at least one secondary case. The shaded areas show the 95% confidence bands clustered on the household level.

**Figure S9:**
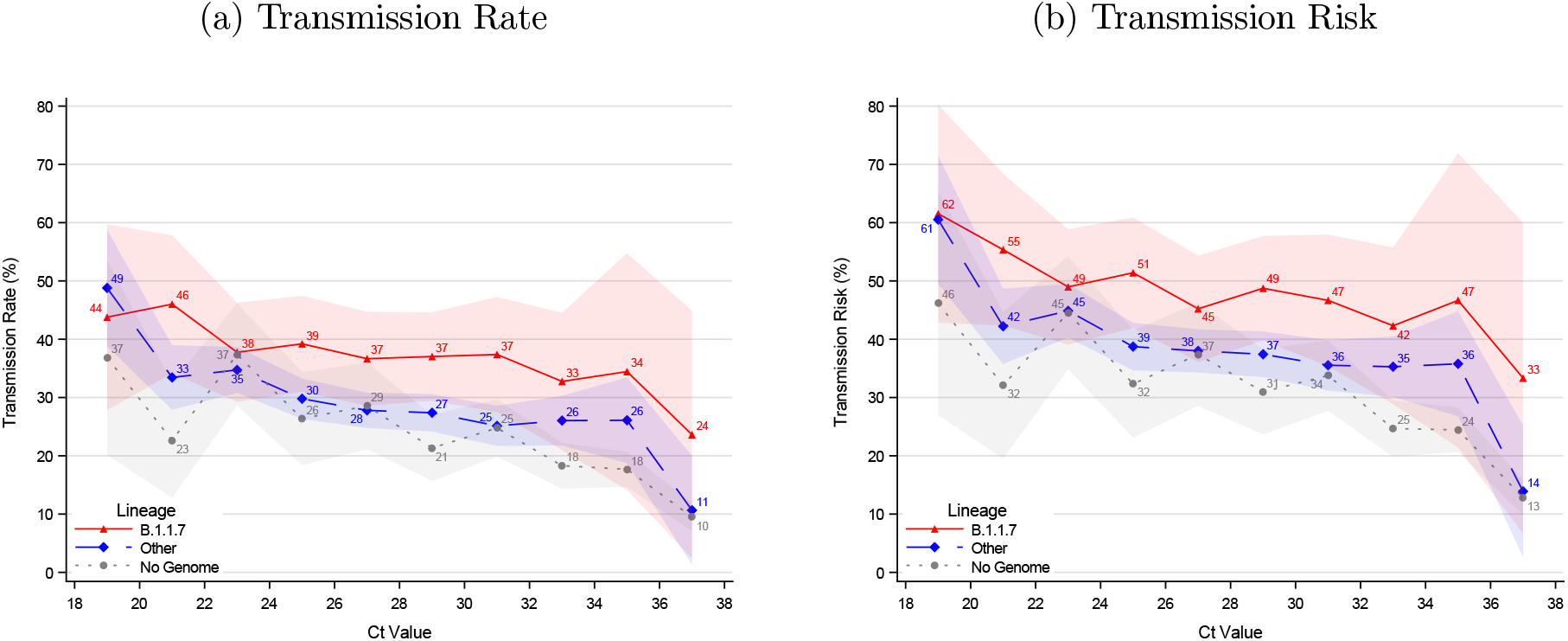
Transmissibility stratified by lineage and Ct values Notes: The transmission rate describes the proportion of potential secondary cases within the household that were infected. The transmission risk describes the proportion of infected primary cases that infected at least one secondary case. The shaded areas show the 95% confidence bands clustered on the household level.

**Table S3:** Sensitivity analysis for the definition of co-primary cases: Odds ratio estimates

|  | **I** | **II** | **III** | **IV** |
| --- | --- | --- | --- | --- |
| **Days for including Sec. Cases** | **1-14** | **2-14** | **3-14** | **4-14** |
| Transmission Rate, B.1.1.7 | 1.62 | 1.61 | 1.59 | 1.60 |
| 95%-CI | (1.39-1.90) | (1.37-1.89) | (1.35-1.88) | (1.34-1.90) |
| Transmission Risk, B.1.1.7 | 1.61 | 1.65 | 1.67 | 1.65 |
| 95%-CI | (1.36-1.90) | (1.38-1.98) | (1.37-2.03) | (1.34-2.04) |
| Constant | ✓ | ✓ | ✓ | ✓ |
| Age, Primary Case | ✓ | ✓ | ✓ | ✓ |
| Ct Value | ✓ | ✓ | ✓ | ✓ |
| Observations | 8,762 | 8,590 | 8,348 | 8,224 |
| Households | 4,172 | 4,033 | 3,847 | 3,761 |

Notes: This table shows sensitivity analysis results for the transmission rate and transmission risk when restricting the inclusion criteria for secondary cases. Column I includes secondary cases that tested positive on days 1-14, i.e., the same as in the paper. Column II only includes secondary cases that tested positive on days 2-14. Column III includes secondary cases that tested positive on days 3-14. Column IV includes secondary cases that tested positive on days 4-14. 95% confidence bands clustered on the household level.

## Appendix C: Statistical Analyses

### Age Structured Transmissibility

To estimate the association between age and transmission rate, stratified by lineage, we estimated the non-parametric regression equation:

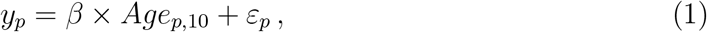

where *Age*_*p*,10_ is the age (in ten-year groups) of the primary case. *β* measures the transmission rate for each ten-year age group of the primary cases. *ε*_*p*_ denotes the error term, clustered on the household (event) level.

### Additive vs. Multiplicative Effect of B.1.1.7 Transmissibility

We wanted to evaluate whether the effect of being infected with B.1.1.7 relative to being infected with other lineages was additive or multiplicative, which is important for designing proper simulation models. With binomial outcomes the canonical link function is the logit function which corresponds to a multiplicative effect. An additive effect of the covariates can be modelled by using the identity link in a generalized linear regression model.

Thus, to estimate the transmissibility effect of B.1.1.7 compared with other lineages, we estimated the model with the following linear predictor:

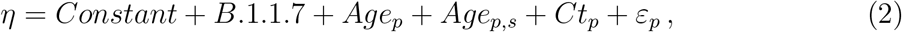

while varying the link function to compare the model fit of an additive versus a multiplicative effect.

As the two models include the same parameters, the model fits can be compared using the Akaike Information Criterion (AIC). Furthermore, reduced versions of the linear predictors were tested. Across all three model specifications and for both transmission rate and transmission risk, we found that the logit model had a lower AIC and, thereby, was a better fit compared with the identity model, implying that the increased transmissibility is multiplicative and not additive (Table S4 and S5).

**Table S4:** Comparison of additive vs. multiplicative effect, Transmission Rate

|  | Linear | Logit | Linear | Logit | Linear | Logit |
| --- | --- | --- | --- | --- | --- | --- |
| AIC | 6,280 | 6,277 | 6,302 | 6,273 | 4,963 | 4,953 |
| B.1.1.7 | ✓ | ✓ | ✓ | ✓ | ✓ | ✓ |
| Constant | ✓ | ✓ | ✓ | ✓ | ✓ | ✓ |
| Age, Primary Case | ✓ | ✓ | ✓ | ✓ | ✓ | ✓ |
| Age, Pot. Sec. Case |  |  | ✓ | ✓ | ✓ | ✓ |
| Ct Value |  |  |  |  | ✓ | ✓ |
| Observations | 10,834 | 10,834 | 10,834 | 10,834 | 8,762 | 8,762 |
| Households | 5,241 | 5,241 | 5,241 | 5,241 | 4,172 | 4,172 |

Notes: This table provides a comparison of an additive and multiplicative model using the Akaike Information Criteria (AIC). The transmission rate describes the proportion of potential secondary cases within the household that were infected.

**Table S5:** Comparison of additive vs. multiplicative effect, Transmission Risk

|  | Linear | Logit | Linear | Logit |
| --- | --- | --- | --- | --- |
| AIC | 6,900 | 6,898 | 5,455 | 5,455 |
| B.1.1.7 | ✓ | ✓ | ✓ | ✓ |
| Constant | ✓ | ✓ | ✓ | ✓ |
| Age, Primary Case | ✓ | ✓ | ✓ | ✓ |
| Ct Value |  |  | ✓ | ✓ |
| Observations | 10,834 | 10,834 | 8,762 | 8,762 |
| Households | 5,241 | 5,241 | 4,172 | 4,172 |

Notes: This table provides a comparison of an additive and multiplicative model using the Akaike Information Criteria (AIC). The transmission risk describes the proportion of infected primary cases that infected at least one secondary case.

We also tested whether other explanatory variables had any significant effect on the increased transmissibility, e.g., household size. Moreover, we investigated the interaction effect, e.g., to see whether the effect was different across age groups. (Data not shown.)

